# Serologic cross-reactivity of SARS-CoV-2 with endemic and seasonal *Betacoronaviruses*

**DOI:** 10.1101/2020.06.22.20137695

**Authors:** Jennifer Hicks, Carleen Klumpp-Thomas, Heather Kalish, Anandakumar Shunmugavel, Jennifer Mehalko, John-Paul Denson, Kelly Snead, Matthew Drew, Kizzmekia Corbett, Barney Graham, Matthew D Hall, Matthew J Memoli, Dominic Esposito, Kaitlyn Sadtler

**Affiliations:** Trans-NIH Shared Resource on Biomedical Engineering and Physical Science, National Institute of Biomedical Imaging and Bioengineering, National Institutes of Health, Bethesda MD 20894; Section on Immuno-Engineering, National Institute of Biomedical Imaging and Bioengineering, National Institutes of Health, Bethesda MD 20892; National Center for Advancing Translational Sciences, National Institutes of Health, Rockville MD, 20850; Protein Expression Laboratory, NCI RAS Initiative, Cancer Research Technology Program, Frederick National Laboratory for Cancer Research, Frederick, MD 21702; Vaccine Research Center, National Institute for Allergy and Infectious Disease, National Institutes of Health, Bethesda, MD 20892; LID Clinical Studies Unit, Laboratory of Infectious Diseases, Division of Intramural Research, National Institute for Allergy and Infectious Disease, National Institutes of Health, Bethesda, MD 20894

**Keywords:** Infectious disease, serology, coronavirus

## Abstract

In order to properly understand the spread of SARS-CoV-2 infection and development of humoral immunity, researchers have evaluated the presence of serum antibodies of people worldwide experiencing the pandemic. These studies rely on the use of recombinant proteins from the viral genome in order to identify serum antibodies that recognize SARS-CoV-2 epitopes. Here, we discuss the cross-reactivity potential of SARS-CoV-2 antibodies with the full spike proteins of four other *Betacoronaviruses* that cause disease in humans, MERS-CoV, SARS-CoV, HCoV-OC43, and HCoV-HKU1. Using enzyme-linked immunosorbent assays (ELISAs), we detected the potential cross-reactivity of antibodies against SARS-CoV-2 towards the four other coronaviruses, with the strongest cross-recognition between SARS-CoV-2 and SARS /MERS-CoV antibodies, as expected based on sequence homology of their respective spike proteins. Further analysis of cross-reactivity could provide informative data that could lead to intelligently designed pan-coronavirus therapeutics or vaccines.

## INTRODUCTION

The SARS-CoV-2 pandemic has reached almost every country on Earth. As with many viral infections, our immune system responds to SARS-CoV-2 infection through a variety of cellular and humoral effectors. These include antibodies produced by B cells, which can be formed against various viral proteins. For SARS-CoV-2, antibodies have been detected that recognize three of the four SARS-CoV-2 proteins exposed on the surface of the viral capsid: the nucleocapsid (N), envelope (E), and spike (S) proteins (1). The spike protein forms as a homotrimer and mediates receptor binding through its receptor binding domain (RBD) to host cell ACE2 and is thus the major target of neutralizing antibody responses (2, 3). When testing for the presence of SARS-CoV-2 antibodies, researchers have utilized the full spike ectodomain as well as the RBD domain alone for antigens in enzyme-linked immunosorbent assays (ELISAs) and other serologic assays (4).

The zoonotic *Betacoronaviruses* SARS-CoV and SARS-CoV-2 (endemic/pandemic B-lineage), and MERS (endemic C-lineage) transferred primarily from bats, while the viruses OC43 and HKU1 (seasonal A-lineage coronaviruses) are endemic in humans (5, 6). All of these viruses bear the spike protein on their surface (7, 8). As such, anti-spike antibodies are common in response to each of the five human-infecting *Betacoronaviruses* (9–11). Knowledge of cross-reactivity of anti-spike antibodies against different viruses is critical for understanding of SARS-CoV-2 immunity of individuals who have had prior exposure to other *Betacoronaviruses* and of potential future immunity of COVID-19 survivors to other coronaviruses (12). Furthermore, knowledge of cross-reactivity is necessary to understand and properly interpret results from serologic studies such as serosurveys and clinical antibody tests (13, 14). Previous research has shown minimal cross-reactivity between RBD domains from differing coronaviruses; however, these studies largely ignore the rest of the spike protein, which will be an important consideration for identification of potential therapeutic antibodies and can be used *in vitro* to help identify polyclonal responses that are not detected with RBD alone (15).

Here, we evaluated the serologic reactivity of pre-pandemic archival blood serum samples (pre-2019) and samples collected in April 2020 from a community highly affected by SARS-CoV-2. Utilizing twelve previously reported ELISAs (15), we tested IgG, IgM and IgA reactivity against spike proteins from SARS-CoV-2, MERS-CoV, SARS-CoV, HCoV-OC43, and HCoV-HKU1 (**Fig. 1**).

**Figure 1:**
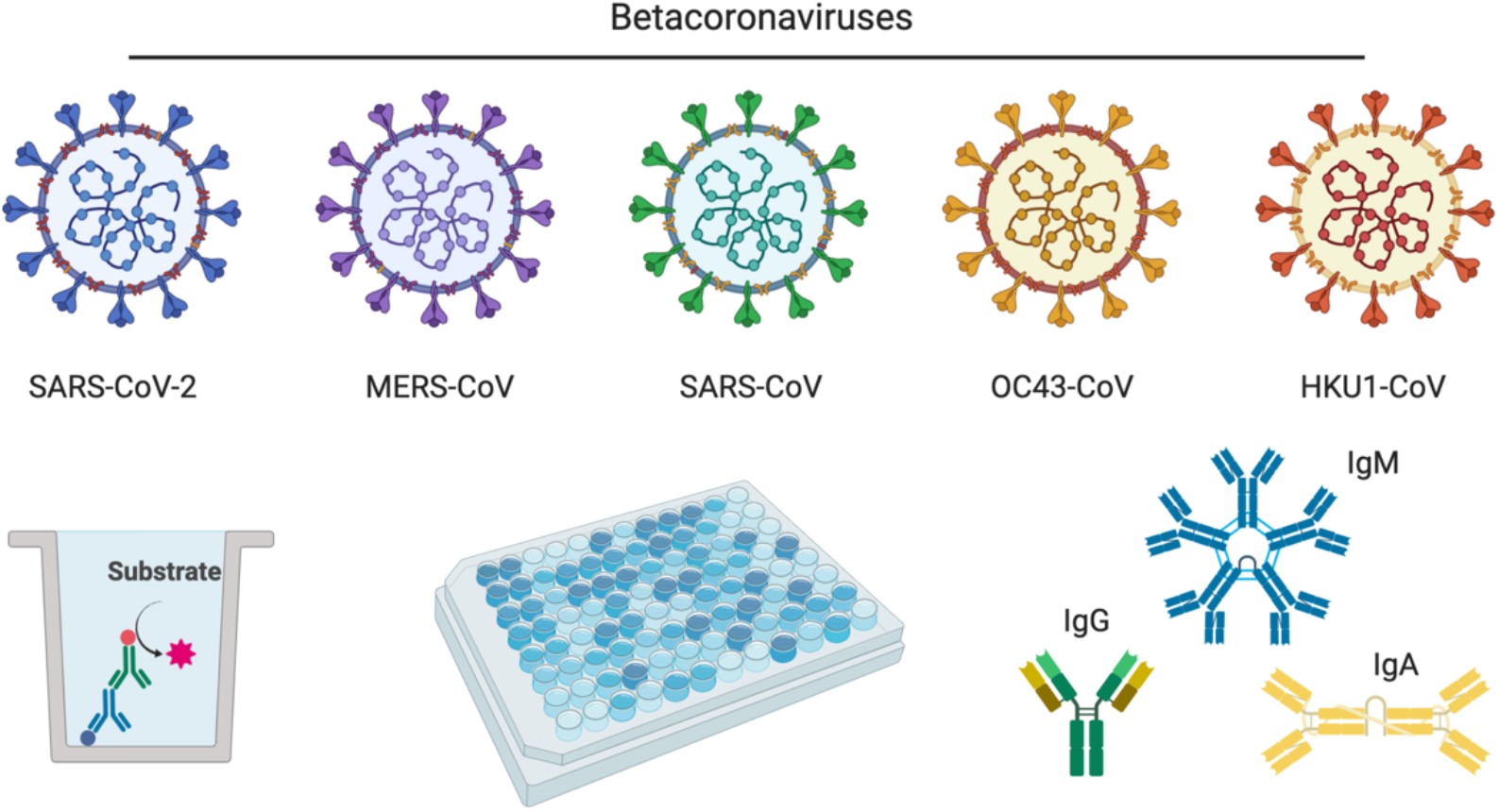
Five different *Betacoronaviruses* with potential for cross-reactivity. We evaluated the serologic cross-reactivity of five *betacoronaviruses* in the context of ELISA-based detection of IgG, IgM, and IgA antibodies against SARS-CoV-2.

## RESULTS

### Sequence homology between pandemic, endemic, and seasonal coronaviruses

To evaluate the potential for cross-reactivity, we first compared the spike protein sequence homology among SARS-CoV-2, MERS-CoV, SARS-CoV, HCoV-OC43, and HCoV-HKU1 (**Fig. 2, Supplementary Figure 1**). The greatest homology was between SARS-CoV-2 and SARS-CoV (76% identity, 87% similarity), followed by MERS (42% identity, 58% similarity) and lastly OC43/HKU1 (OC43: 30% identity, 41% similarity; HKU1: 29% identity, 40% similarity). A-lineage OC43 and HKU1 are more similar to each other (64% identity, 75% similarity) than to the two endemic *Betacoronaviruses*. There is a larger fraction of homology towards the C-terminus of the protein in all coronavirus spike proteins, which represents the major structural regions of the protein including the heptad repeat regions responsible for insertion of the fusion peptide into the host cell membrane. Homology is significantly lower in the N-terminal regions of spike, with significant lack of similarity in the regions including the receptor-binding domain, correlating with the difference in receptors and determinants used for host cell entry in the different *Betacoronaviruses* (MERS-CoV: receptor dipeptidyl peptidase-4 (DPP4), SARS-CoV/SAR-CoV-2: ACE2, OC43/HKU1: the sugar N-Acetylneuraminic acid)(8).

**Figure 2:**
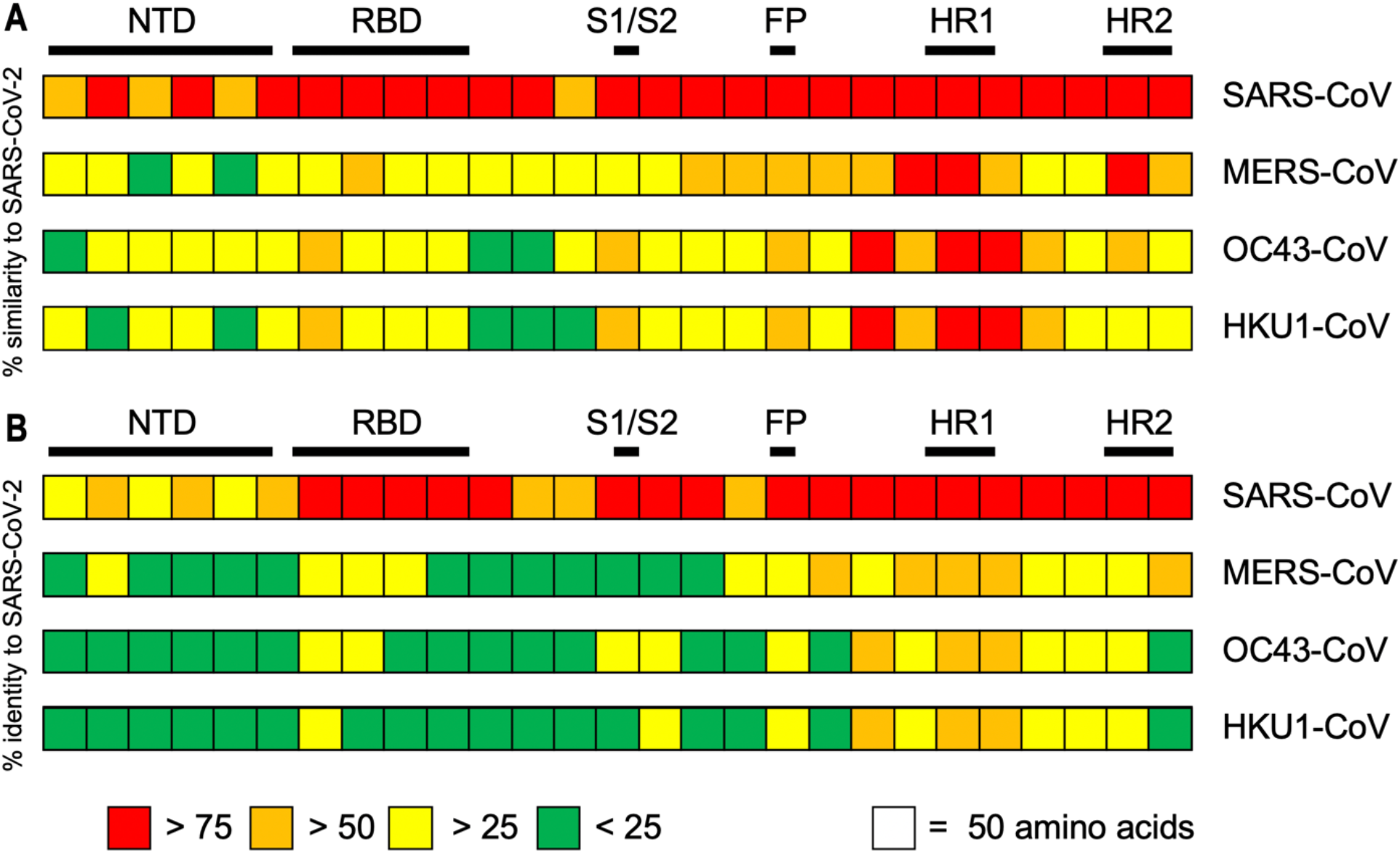
Sequence homology of SARS-CoV-2 with endemic and seasonal *Betacoronaviruses*. SARS-CoV-2 spike ELISA antigen protein sequence aligned with MERS-CoV (MERS), SARS-CoV (SARS1), OC43, and HKU1 *Betacoronaviruses*. (A) Percent (%) similarity to SARS-CoV-2. (B) Percent (%) identity to SARS-CoV-2.

### Serologic reactivity of anti-spike IgG, IgM and IgA antibodies

Functional cross-reactivity was determined through the use of enzyme-linked immunosorbent assays (ELISAs) measuring IgG, IgM and IgA subclasses, representing mature, early stage, and mucosal specific serologic responses, respectively. We produced recombinant soluble spike proteins of SARS-CoV-2, MERS, SARS-CoV, OC43, and HKU1 using the Expi293 expression system, which yielded pure, intact ectodomain trimers suitable for ELISA (16). Notably, the yields of all coronavirus spike proteins were significantly different even though all four of five were cloned in identical vectors and contained the same modifications to the wildtype sequences (elimination of furin cleavage site, prefusion-stabilizing proline mutations (2P), similar C-terminal tags), none of which is expected to alter serologic recognition due to their internal locations. The HCoV-OC43 construct has all of these features but the wild-type furin cleavage site is present. Using similar expression conditions, SARS-CoV-2 spike was produced at a maximum of 2 mg/L culture, while the other spike proteins were significantly easier to produce with yields of 5, 11, 8, and 6 mg/L respectively for SARS-CoV, MERS, OC43, and HKU1. We utilized a semi-automated ELISA protocol to detect serum antibodies from pre-2019 archival samples and samples from a community with high SARS-CoV-2 prevalence during the 2020 pandemic (**Fig. 3**). In serum samples collected from healthy volunteers prior to 2019, there was minimal reactivity with SARS-CoV-2, MERS and SARS-CoV. The majority of tested samples (*n* = 114) displayed high IgG reactivity with OC43 and HKU1 spike proteins, consistent with the extensive spread of seasonal *Betacoronavirus* infections within the United States (**Fig. 3a**,**b**). As reported previously, we detected a high proportion of donors who seroconverted and were SARS-CoV-2 IgG+ in a community in New York City, along with a significant number of IgM and IgA seropositive donors, including several donors who were non-symptomatic (15). All samples had low levels of IgM reactivity against MERS, SARS-CoV, OC43, and HKU1 (**Fig. 3c**,**d**). IgA antibodies were present at higher levels than IgM, but still well below levels of IgG, correlating well with biologic prevalence of antibody classes in response to pathogens (**Fig. 3e**,**f**).

**Figure 3:**
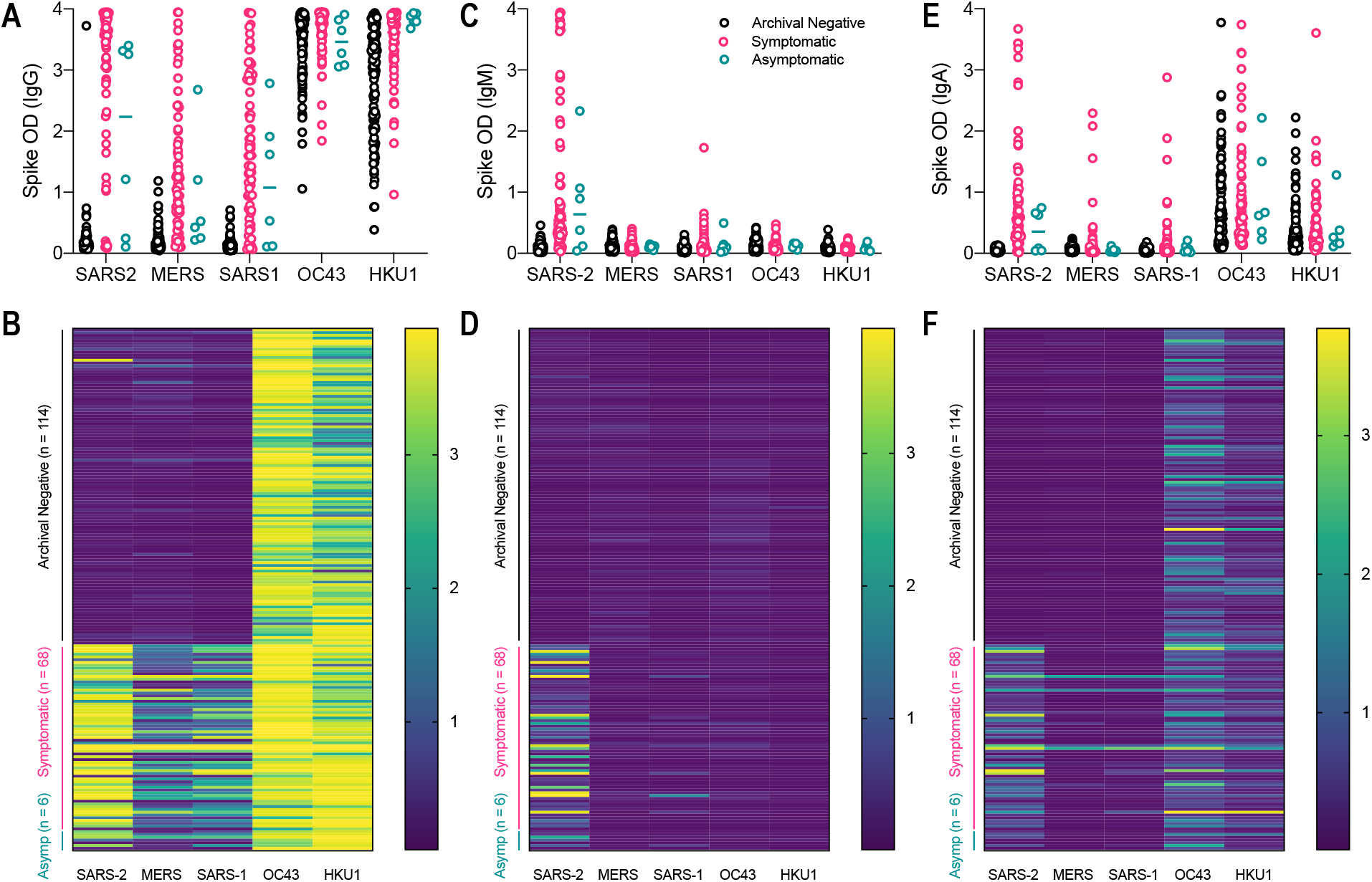
Serologic positivity of immunoglobulins G, M and A for five *Betacoronaviruses* in pre-2019 and high prevalence SARS-CoV-2 blood donors. Signal intensity in archival negative (pre-2019, black), hot-spot community symptomatic (pink), and hot-spot community asymptomatic (teal) blood donors for (a-b) IgG, (c-d) IgM, and (e-f) IgA.

### Minimal linear correlation of SARS-CoV-2 signal intensity with other Betacoronaviruses

When comparing the assay absorbance signal (optical density, OD) between SARS-CoV-2 and the other spike proteins in the high-incidence population, we saw a stronger correlation of signal intensity between SARS-CoV-2 and SARS-CoV IgG (Correlation = 0.711, R^2^ = 0.505) and the lowest correlation with HKU1 (Correlation = 0.281, R^2^ = 0.079) (**Fig. 4a, Supplementary Figure 2**). Though there was not a precise linear correlation for IgG, donors who represented signal intensity in the lower 50% of SARS-CoV-2 absorbance readings did have a significantly lower MERS and SARS-CoV signal intensity when compared to the upper 50% of SARS-CoV-2 intensity (**Fig. 5**). Overall, these data suggest some cross-reactivity occurs that is more easily detectable at high titers of antibody.

**Figure 4:**
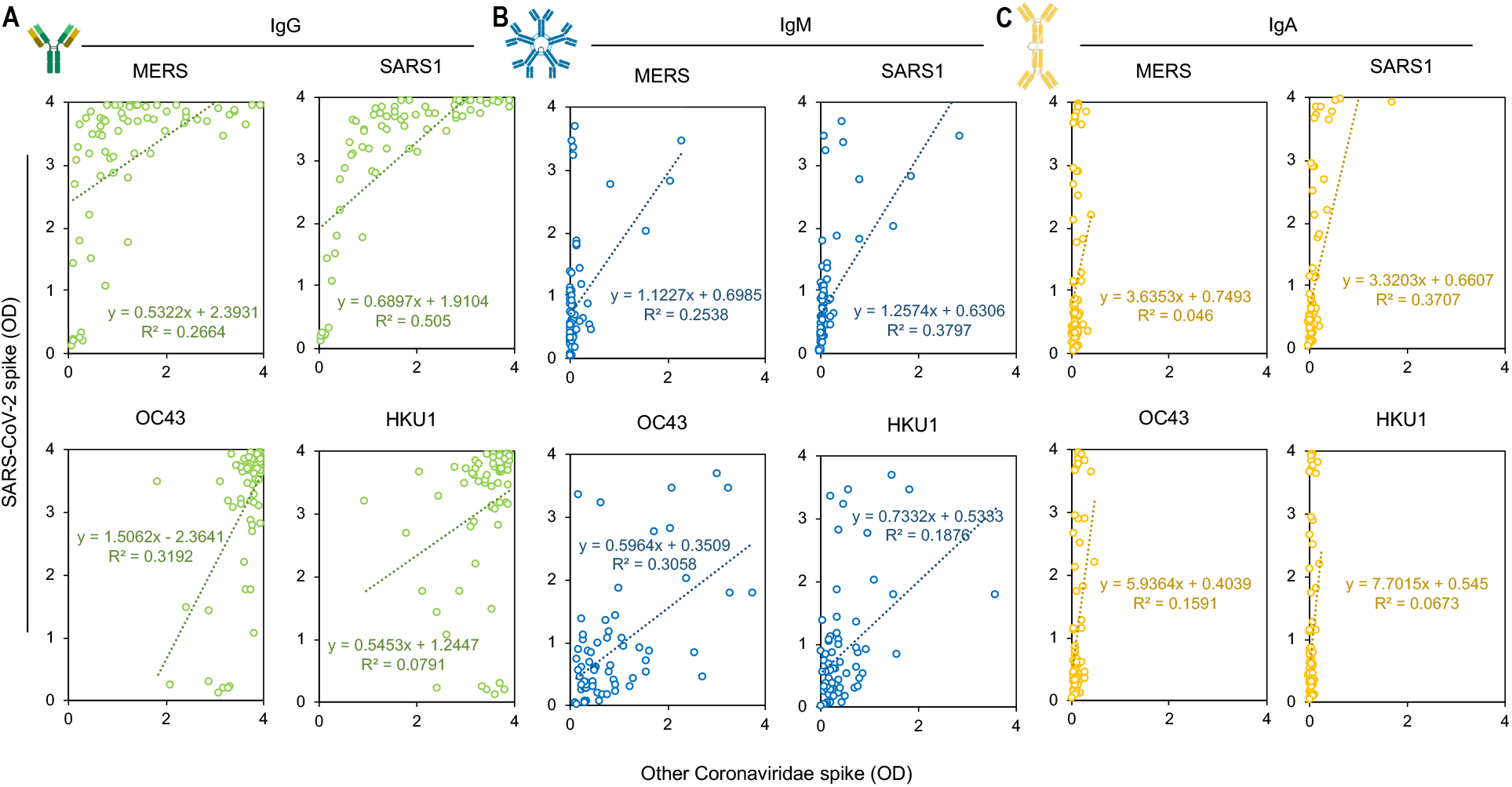
SARS-CoV-2 signal intensity compared with signal intensity of other *Betacoronaviruses* in pandemic hot-spot community blood draws. (a) Anti-spike IgG signal intensity (b) Anti-spike IgM signal intensity, and (c) Anti-spike IgA signal intensity.

**Figure 5:**
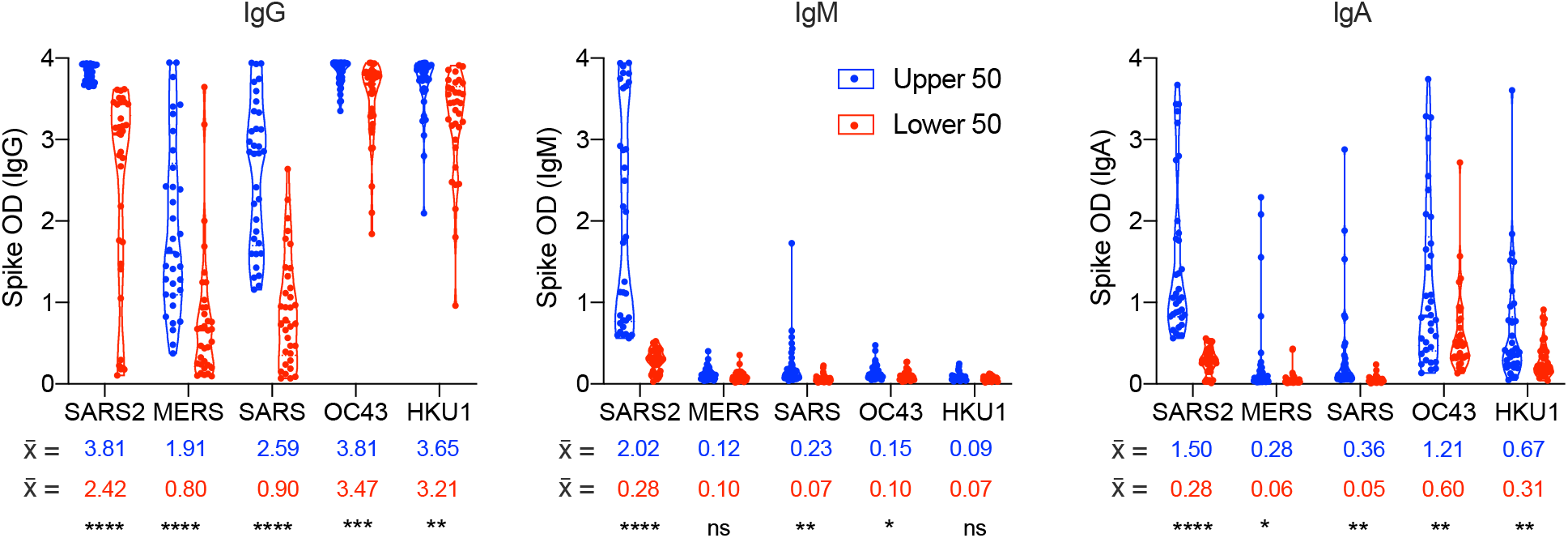
High titers of SARS-CoV-2 spike antibodies correlate with an increase in ELISA signal intensity for other *Betacoronavirus* reactivity. Comparison of the mean absorbance (optical density, OD) of the upper (blue) and lower (red) 50% of SARS-CoV-2 signal intensity for (a) IgG, (b) IgM, and (c) IgA.

### Cross-reactivity of SARS-CoV-2 IgG antibodies with endemic and seasonal coronaviruses

Since we observed a difference in the IgG signal intensity of other *Betacoronaviruses* with high levels of SARS-CoV-2 antibodies, we further analyzed the relationship between SARS-CoV-2 seroprevalence and antibody titer with SARS-CoV, MERS, OC43, and HKU1 in pre-pandemic (pre-2019), high-prevalence symptomatic donors, and high-prevalence asymptomatic donors (**Fig. 6, Supplementary Figure 3**). Overall, archival pre-2019 samples displayed an equivalent low signal intensity of SARS-CoV-2, MERS, and SARS-CoV spike reactivity (**Fig. 6a**). One cross-reactive donor from this group was negative for both MERS and SARS-CoV. As previously discussed, the majority of donors were OC43 and HKU1 seropositive due to the broad circulation of these viruses in humans. In the high incidence community, for both symptomatic and asymptomatic individuals, there appeared to be a correlation in SARS-CoV-2 signal intensity with MERS and SARS-CoV. To further analyze this, we directly compared the signal intensity of archival sample controls to the high-incidence pandemic population (**Fig. 6b**). There was a significant difference in signal intensity of MERS, SARS-CoV, OC43, and HKU1, suggesting potential cross-reactivity of SARS-CoV-2 IgG antibodies with MERS, SARS-CoV, OC43 and HKU1 spike proteins.

**Figure 6:**
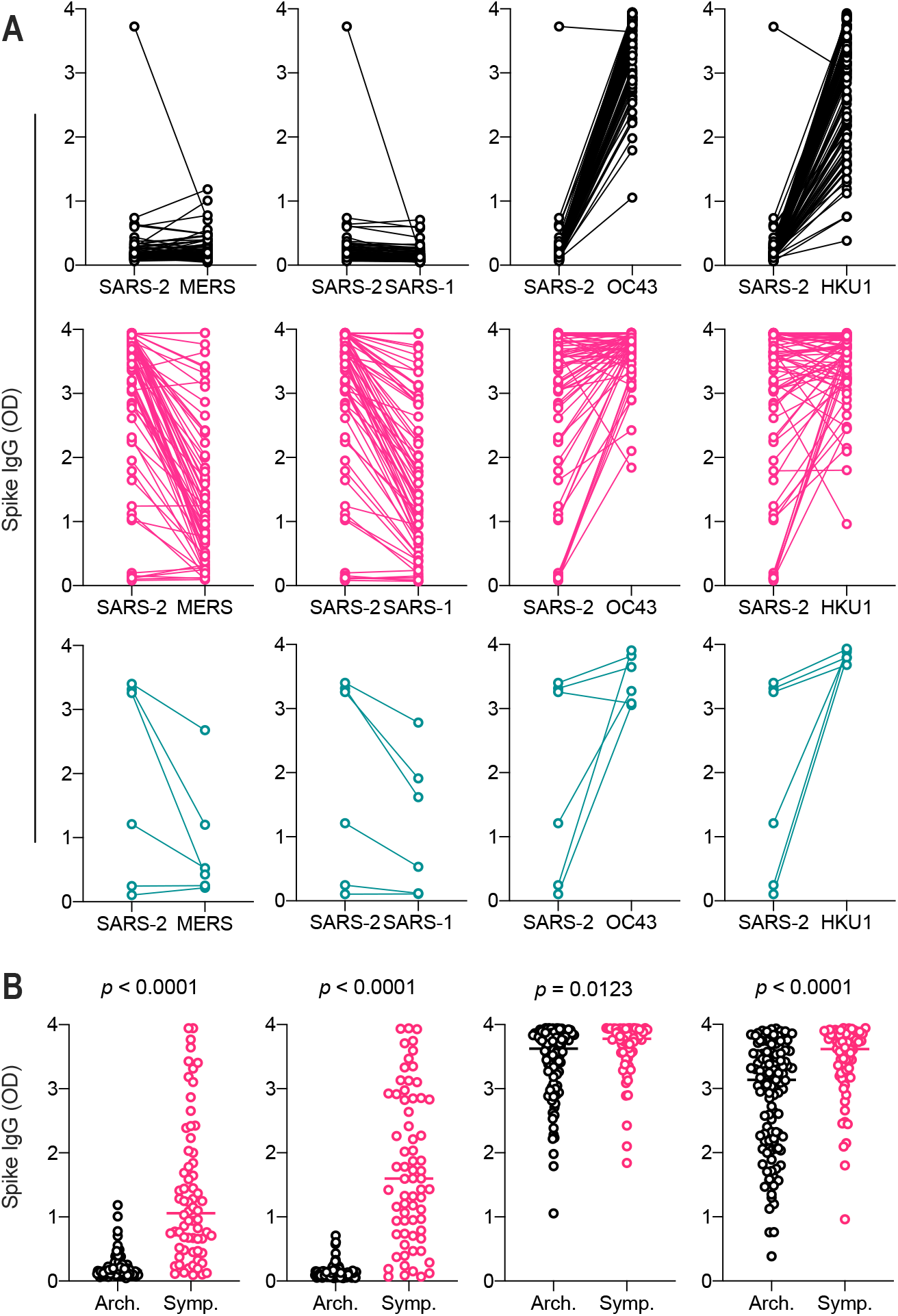
Anti-spike IgG signal intensity in SARS-CoV-2 seropositive and seronegative blood samples. (a) Relationship of SARS-CoV-2 spike IgG signal intensity in archival (black), symptomatic high exposure (pink) and asymptomatic high exposure (teal) donors. (b) Comparison of archival sample IgG reactivity with symptomatic high exposure sample reactivity. Students T-test.

## DISCUSSION

Cross-reactivity of antibodies with multiple coronaviruses is an important consideration in studying the SARS-CoV-2 pandemic, both technically, for identifying individuals who have been exposed to and recovered from the virus, as well as therapeutically, to identify broadly neutralizing antibodies or epitopes on multiple coronavirus subtypes (12, 17, 18). Accordingly, we analyzed potential serologic cross-reactivity of antibodies with spike proteins derived from SARS-CoV-2 as well as two endemic (MERS, SARS-CoV) and two seasonal (OC43, HKU1) *Betacoronavirus* species. It is unclear, in terms of plasmid-based protein expression, why there is so much variability in spike protein expression levels between the different viruses, but this argues again for significant differences in the behavior of these proteins regardless of their primary sequence homology.

Antibodies that react with the spike proteins of OC43 and HKU1 are highly prevalent in the general population of the United States as determined by their measurement in archival pre-2019 serum samples. Previous reports of their prevalence show that the majority of children are exposed to OC43 and seroconvert early in life (19). The detection of high serologic reactivity of archival controls with HKU1 might, thus, be due to the strong seroprevalence of OC43 antibodies. Further studies would be needed to determine this interaction, though due to the high level of sequence and structural homology of their spike proteins, such a cross-reactivity between the two tested seasonal *Betacoronaviruses* would not be surprising.

When compared to reactivity with the SARS-CoV-2 spike protein, antibodies that react to OC43 and HKU1 have minimal cross-reactivity with the pandemic SARS-CoV-2 or two other endemic coronaviruses, MERS and SARS-CoV. This phenotype correlates with the sequence homology of these proteins, wherein SARS-CoV-2 spike is more similar to SARS-CoV and MERS, as opposed to OC43 and HKU1 seasonal coronaviruses.

When comparing serum from healthy volunteers collected pre-2019 (archival controls) to those from a high-exposure community, we observe that SARS-CoV-2 antibodies react intermediately with MERS and SARS-CoV spike proteins. The mean ELISA signal intensity is significantly greater for both MERS and SARS-CoV when comparing archival controls versus the high-incidence community. Although there is minimal linear correlation between signal intensity of SARS-CoV-2 and MERS/SARS-CoV, the higher titer SARS-CoV-2 donors also display a significantly higher MERS and SARS-CoV signal intensity compared to their lower titer counterparts within the same population.

Given the low seroprevalence of SARS-CoV and MERS outside of their endemic regions, and the significantly lower reactivity of SARS-CoV-2 patient sera to SARS-CoV and MERS spike proteins, it is likely that any reactivity between the pandemic SARS-CoV-2 pandemic and MERS/SARS-CoV endemic viruses would result in minimal noise between SARS-CoV-2 signal and endemic coronavirus signal in serological assays. In countries with a higher prevalence of MERS & SARS-CoV, researchers should include thorough analysis of archival patient sera (pre-2019), including sera from known SARS-CoV and MERS convalescent patients, to properly analyze the resulting data and adjust any estimates of seropositivity as needed. No clinical serology studies of SARS-CoV-2 immunity in populations previously infected with either SARS or MERS have yet emerged.

Additionally, individuals who have strongly seroconverted after SARS-CoV-2 infection, and who display cross-reactivity for both MERS and SARS-CoV spike proteins, are of great interest for translational study. These individuals could potentially harbor antibodies that are universally reactive to multiple *Betacoronaviruses* and, if these antibodies are functional for neutralization, could be important to identify to inform the development of novel therapeutics or vaccines.

## MATERIALS & METHODS

### Human serum samples

Archival (pre-2019) serum samples (*n* = 114) were collected between January 2014 and December 2018 from healthy adults (aged 18 – 55 years) through an existing NIH study NCT01386424. High-incidence community samples are deidentified uncoded samples donated from a community blood draw from donors in New York and New Jersey in April 2020. Twenty-two (22) of these donors had a previous SARS-CoV-2 nasopharyngeal swab PCR-based diagnosis, 46 were symptomatic but undiagnosed, and 6 were asymptomatic but had known exposure (*n* = 68 symptomatic, *n* = 6 asymptomatic). All clinical trials were conducted in accordance with the provisions of the Declaration of Helsinki and Good Clinical Practice guidelines. All clinical trial participants signed written informed consent prior to enrollment.

### Plasmid sourcing and preparation

SARS-CoV-2, MERS-CoV, HCoV-HKU1 and SARS-CoV spike plasmids were produced from the McLellan lab at UT Austin and NIAID VRC and prepared as previously described (2, 20, 21). Briefly, for HCoV-OC43 S, a mammalian-codon-optimized gene encoding HCoV-OC43 S (GI: 744516696) ectodomain with a C-terminal T4 fibritin trimerization domain, an HRV3C cleavage site, an 8xHis-tag and a Twin-Strep-tag were synthesized and subcloned into the eukaryotic-expression vector pαH. The S1/S2 furin-recognition site was mutated to produce a single-chain S protein and 2 prolines were substituted, following previous-published prefusion stabilizing mutation strategy.

### Protein production and purification

Soluble spike trimers were produced by expression in Expi293 cells and purified by a combination of tangential flow filtration, immobilized metal affinity chromatography, and desalting, following the procedures noted in Esposito et al. Expression was carried out at 37°C for 72 hours prior to harvest. Final purified proteins were validated by a combination of SDS-PAGE and analytical size exclusion chromatography (AnSEC). All spike proteins produced single peaks on AnSEC over a Superdex 200 column, and the peak elution was consistent with the size of a trimeric spike protein. Of note, the OC43 spike protein undergoes cleavage during SDS-PAGE leading to the appearance of two bands at 80 and 100 kDa as well as the appropriately full-length band migrating at 180 kDa. AnSEC confirms that this is an artifact of the SDS-PAGE process, as the protein elutes in a single trimeric peak of the appropriate size.

### Enzyme-linked Immunosorbent Assays

We performed ELISAs as previously described (15). Briefly, spike proteins were suspended at 1 ug/ml in 1x PBS. One hundred (100) microliters of protein suspension was added to each well of a 96-well Nunc MaxiSorp ELISA plate and allowed to coat overnight at 4°C for 16 hours. Wells were washed three times with 300 ul of 1x PBS + 0.05% Tween20 (wash buffer) followed by blocking for 2 hours at room temperature with 200 ul of 1x PBS + 0.05% Tween20 + 5% Non-fat dry milk (blocking buffer). Wells were washed again three times with 300 ul of wash buffer prior to addition of 100 ul of sample diluted in blocking buffer (serum samples were heat inactivated for 45 minutes at 56°C and diluted at 1:400 in blocking buffer). Samples were incubated for 1 hour at room temperature, then washed three times with 300 ul of wash buffer. One hundred (100) microliters of 1-Step Ultra TMB Substrate (ThermoFisher) was added and the plate was incubated for 10 minutes prior to stopping the reaction with 1N sulfuric acid (Stop Solution, ThermoFisher). Absorbance was read at 450 nm and 650 nm on a BioTek Epoch2 plate reader. The process is semi-automated through the use of a BioTek EL406 plate washer/dispenser and two BioStack 4 plate stackers to minimize plate-to-plate variation and increase throughput (see Klumpp-Thomas C, Kalish H et al. 2020 for detailed automation methods).

### Data Analysis

Absorbance values (optical density) were collected at 450 and 650 nm. A650 was subtracted from A450 to remove background signal. Data were subsequently analyzed utilizing Microsoft Excel and GraphPad Prism.

## Data Availability

Data are available upon request after formal publication of manuscript.

## ACKNOWLEDGEMENTS

The authors would like to thank Golan Ben-Oni, Rabbi Shua Brook, Dr. Adam Polinger, Dr. Avi Rosenberg, and the Jewish community of New York and New Jersey for their generous donation of blood samples use in this assay. We thank members of the FNLCR Protein Expression Laboratory (William Gillette, Simon Messing, and Vanessa Wall) for support in DNA production and protein purification. This research was supported in part by the Intramural Research Program of the NIH, including the National Institute of Biomedical Imaging and Bioengineering, the National Institute of Allergy and Infectious Disease, and the National Center for Advancing Translational Sciences. This project has been funded in part with Federal funds from the National Cancer Institute, National Institutes of Health, under contract number HHSN261200800001E.

## Disclaimer

The NIH, its officers, and employees do not recommend or endorse any company, product, or service.

## SUPPLEMENTAL FIGURES AND LEGENDS

**Supplementary Figure 1:**
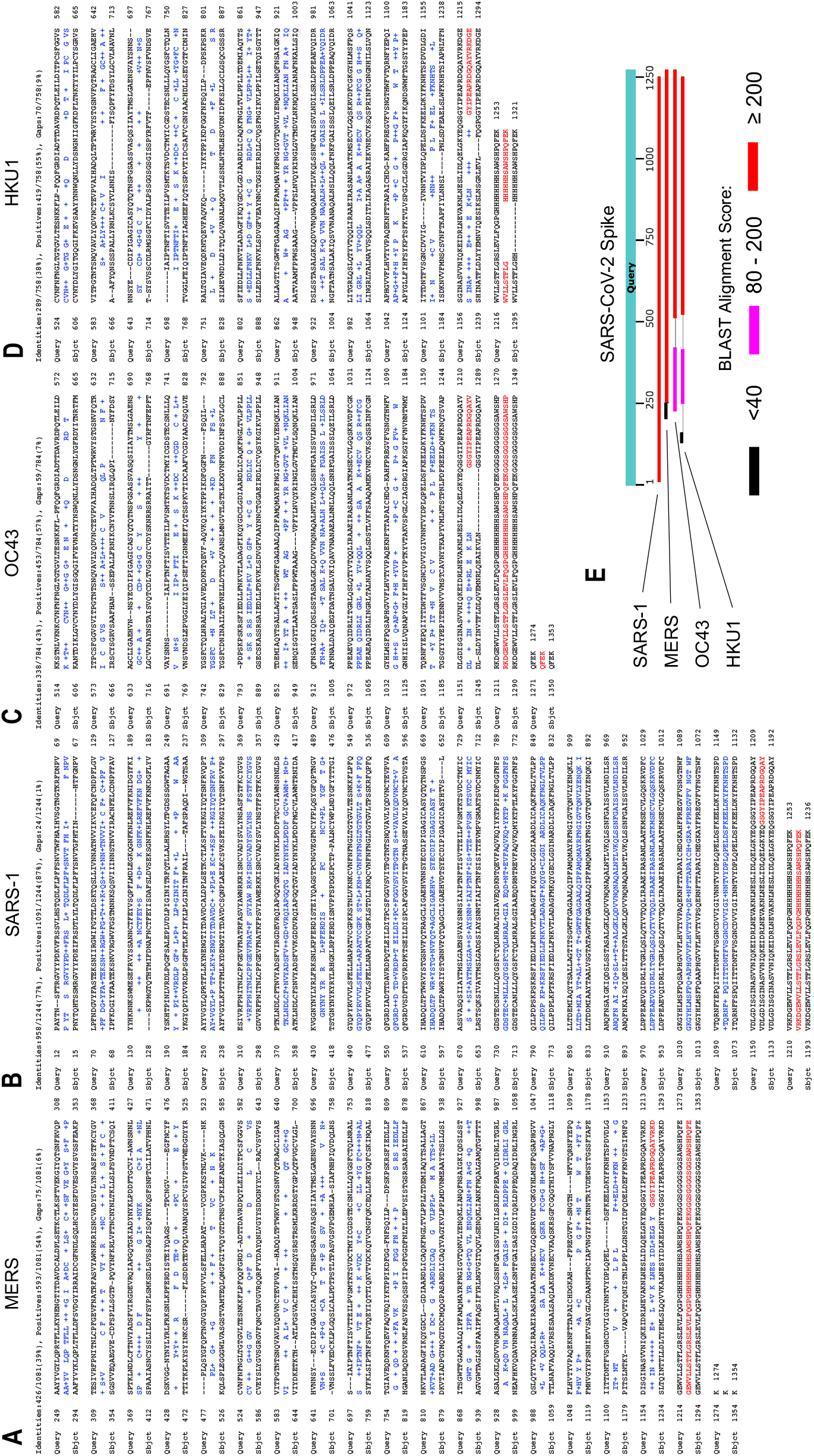
BLAST alignment of 4 coronaviruses with SARS-CoV-2.

**Supplementary Figure 2:**
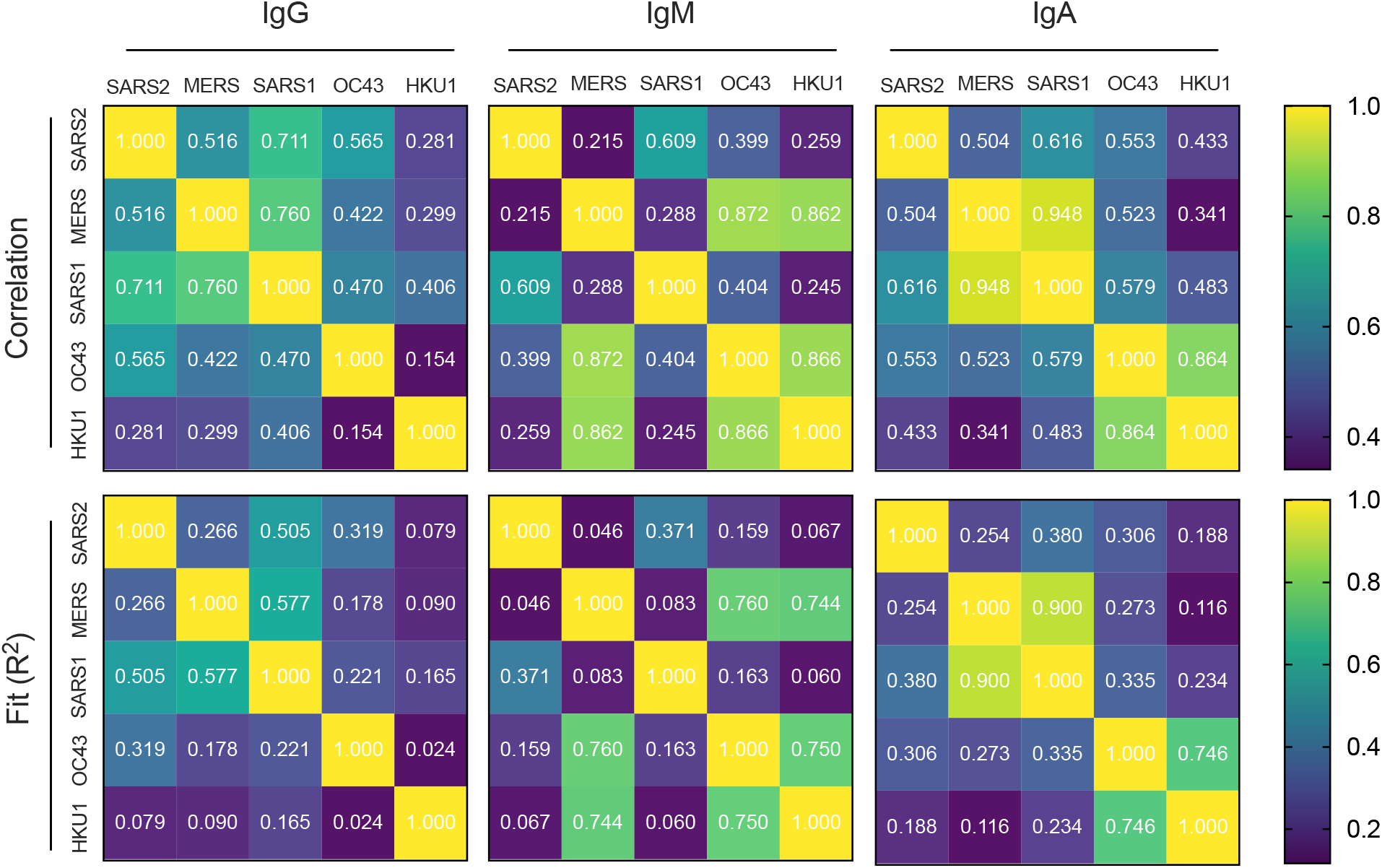
Linear Correlation Statistics of *Betacoronaviruses*.

**Supplementary Figure 3:**
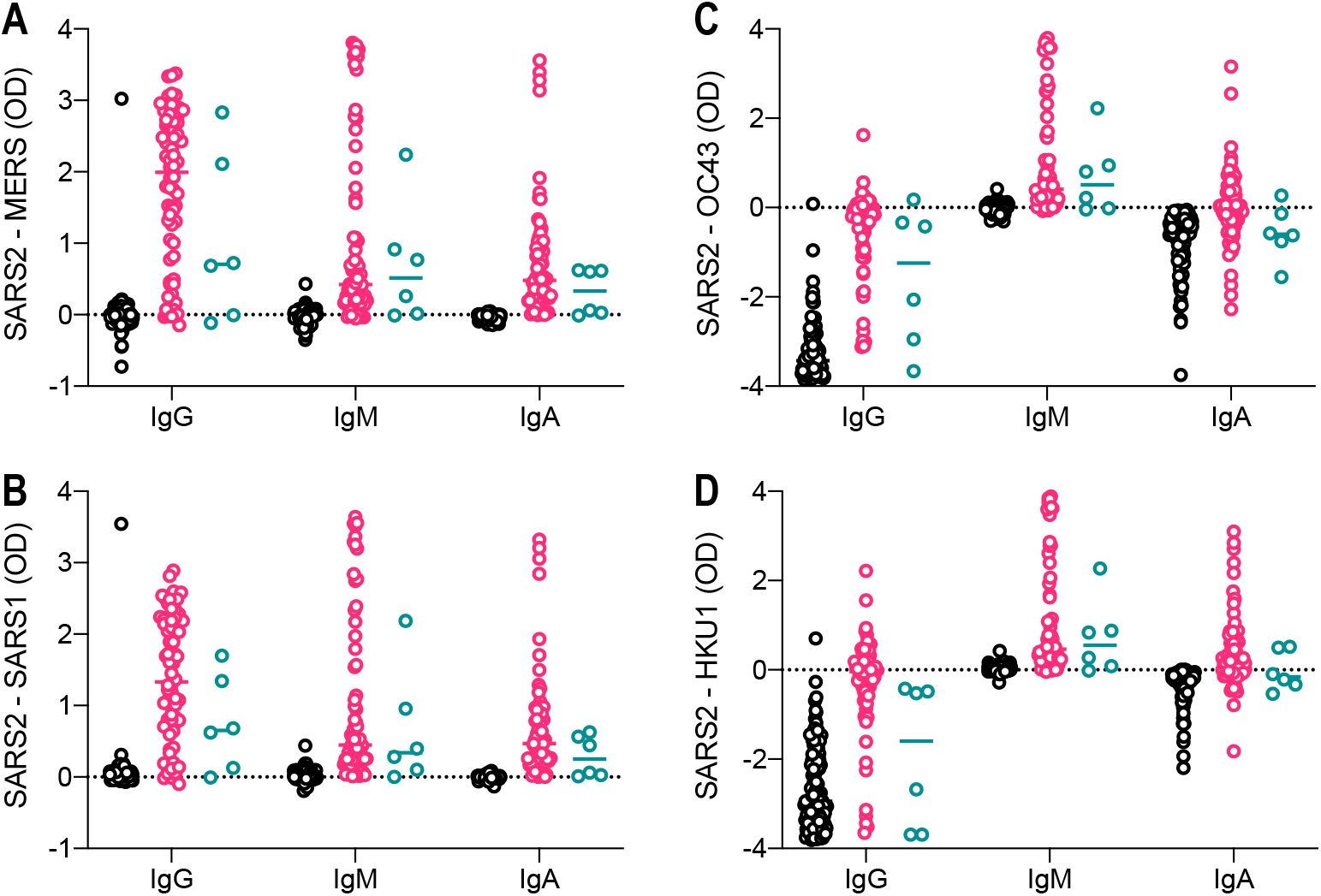
Differential signal intensity of SARS-CoV-2 spike with other *Betacoronaviruses*. Signal intensity displayed as SARS-CoV-2 absorbance (A450 – A650) minus signal intensity of other *Betacoronaviruses*. (a) MERS, (b) SARS-CoV, (c) OC43, and (d) HKU1. Archival negative = black, pandemic hot-spot symptomatic = pink, pandemic hot-spot asymptomatic = teal. n =114 archival negative, n = 68 hot-spot symptomatic, n = 6 hot-spot asymptomatic.

